# Effects of theta burst stimulation on neural connectivity and visual perception following attention modification of own-face viewing in body dysmorphic disorder

**DOI:** 10.64898/2026.05.25.26354053

**Authors:** Joel P. Diaz-Fong, Hayden J. Peel, Kaixi Zhang, Jessica Qian, Madison Lewis, Wan-Wa Wong, Andrew F. Leuchter, Reza Tadayonnejad, Daphne Voineskos, Gerasimos Konstantinou, Eileen Lam, Daniel M. Blumberger, Jamie D. Feusner

## Abstract

**Background:** Individuals with body dysmorphic disorder (BDD) misperceive defects in their appearance. Evidence suggests that visual processing abnormalities may underlie this core symptom. Previous studies testing perceptual and attentional interventions and non-invasive neuromodulation suggest these abnormalities may be modifiable, but their combined effects on neural connectivity and perceptual processing remain unclear.

**Methods:** Unmedicated participants with BDD or subclinical BDD received one session each of intermittent and continuous theta burst stimulation (TBS) targeting the lateral parietal cortex, combined with a visual attention modification paradigm during functional magnetic resonance imaging, in a crossover design. Dynamic effective connectivity within dorsal and ventral visual stream pathways was calculated, and global visual processing biases were assessed using the face inversion effect before and after each session.

**Results:** The analytic sample comprised 38 participants (60.5% female). Intermittent TBS increased connectivity in higher-level dorsal pathways during naturalistic viewing following attention modification (β = 0.079, *p* = .008), whereas continuous TBS reduced connectivity in lower-level dorsal pathways (β = −0.118, *p* < .001) and increased connectivity in ventral pathways (higher: β = 0.239, p < .001; lower: β = 0.079, *p* = .014). These changes were accompanied by differential effects on global visual processing.

**Conclusions:** Combined neuromodulation and visual attention modification modulate visual system connectivity and perceptual processing in individuals with BDD symptoms. These findings support a mechanistic link between dorsal–ventral stream dynamics and perceptual biases. Integrating neuromodulation with perceptual retraining may represent a viable approach for targeting distorted appearance perception.

ClinicalTrials.gov: NCT05607121

## Introduction

Body dysmorphic disorder (BDD) is a debilitating psychiatric condition in which individuals misperceive aspects of their appearance as conspicuously flawed or defective, despite these features appearing unnoticeable or minuscule to others (1). Affecting approximately 2% of the general population (2–5) and up to 20% of individuals presenting for cosmetic surgery (6,7), BDD is associated with substantial functional impairment and an elevated risk for suicide (8). Although concerns may involve any appearance feature, they most commonly center on the face and head, such as skin, hair, and nose (9). Despite its prevalence and severity, few neurobiological or treatment studies have been conducted, and no treatment has been proven to remediate perceptual distortions of appearance in BDD.

Mechanisms that may underlie the perceptual distortions experienced by individuals with BDD include abnormalities in visual processing systems (10–14). Further, those with BDD exhibit attentional biases; eye-tracking studies have revealed biases away from features they rate as attractive (15) and towards unattractive features (16) and imagined defects (17). These abnormalities have contributed to a model of diminished global/holistic processing and enhanced local/detailed processing (18–21), attributed to “bottom-up” and “top-down” disturbances in perception. Consistent with this model, individuals with BDD exhibit reduced activation and connectivity within the dorsal visual stream and (although less consistently) increased activation and connectivity within the ventral visual stream compared to healthy controls, implicating regions associated with global/holistic visual processing and local/detailed visual processing, respectively (20,21). Because the lateral parietal cortex, including the intraparietal sulcus and superior parietal lobule, is part of the dorsal visual stream and serves as a central hub supporting spatial attention and global visual integration (22,23), it represents a rational neuromodulation target for modifying aberrant visual processing in BDD and was selected as the primary stimulation target in the present study.

Such disturbances in perceptual processing have been observed through psychophysical tests, such as the face inversion effect, a robust phenomenon marked by a reduced recognition accuracy and increased response time when viewing inverted compared to upright faces (24,25). Inversion is thought to disrupt the habitual global strategies used to identify human faces, requiring detailed feature-based processing instead (26). Diminished inversion effects have been observed in individuals with high body dysmorphic concern and in individuals with BDD in response to face (12,13,27) and body (28) stimuli. Thus, while global/holistic processing strategies may be habitually under-used in individuals with BDD and those with high body image concerns, these abnormalities may be modifiable under certain circumstances.

Modification to the visual processing systems of individuals with BDD and healthy controls may be achieved with a visual attention modification paradigm (29). In this paradigm, individuals view photographs of their own face first as they normally would (naturalistically), then while focusing their attention on a translucent crosshair in the center of the photos (ModV), and then naturalistically once again. ModV may reduce the extensive scanpaths observed in individuals with BDD (30) and affect both top-down and bottom-up mechanisms in a two-fold manner, by voluntarily directing attention away from perceived defects towards the central crosshair and by reducing foveal input to those features in favor of peripheral input, which is tuned toward the lower spatial frequencies associated with global, dorsal-stream processing (31), potentially enhancing dorsal visual stream functioning.

Similar modifications to dynamic effective connectivity within these systems have been achieved with theta burst stimulation (TBS), a type of repetitive transcranial magnetic stimulation, in a pilot study of unmedicated adults with BDD with face concerns (32). During naturalistic own-face viewing, dynamic effective connectivity in the anterior dorsal visual stream was increased with high intensity (100% active motor threshold) compared to low intensity stimulation (10% active motor threshold), providing preliminary evidence that exogenous neuromodulation can modify aberrant visual processing mechanisms in individuals with BDD.

Combining exogenous modulation with behavioral attentional modification could induce the functional changes necessary for clinically meaningful improvements in perceptual experiences in those with BDD. To further test the modifiability of aberrant visual processing mechanisms in BDD, the current study combines ModV with TBS. The approach was hypothesized to exert effects through alterations in neuroplasticity induced by exogenous modulation (33), which then enhance the effects and/or durability of attentional modification, or through additive effects of TBS and attentional modification, which have independently been shown to enhance dorsal visual stream function (29,32). Excitatory and inhibitory effects in certain brain regions have traditionally been observed following 600 pulses of intermittent (iTBS) and continuous TBS (cTBS), respectively (34). At this duration, iTBS is believed to induce long-term potentiation-like plasticity through NMDA receptor-dependent Ca²⁺ influx at glutamatergic synapses, while cTBS is thought to induce long-term depression-like plasticity via interneuronal GABAergic inhibition (35). Enhanced dorsal visual stream connectivity following iTBS, as observed in our pilot study, may therefore reflect glutamatergic potentiation at the lateral parietal cortex propagating through downstream visual circuitry.

Using a within-subject crossover design, we examined whether iTBS and cTBS “enhance” and “inhibit”, respectively, the effects of behavioral visual attention modification on brain connectivity and global visual processing in individuals with clinical and subclinical BDD. We hypothesized that iTBS would increase connectivity within the dorsal visual stream and reduce connectivity within the ventral visual stream, whereas cTBS would produce the opposite pattern. We also hypothesized that iTBS would be associated with increased and cTBS with decreased global visual processing biases, measured with the face inversion effect.

## Methods and Materials

### Participants

The study was approved by the Research Ethics Boards at the Centre for Addiction and Mental Health and the University Health Network and was registered with ClinicalTrials.gov (NCT05607121). The study design, methodology, and hypotheses were pre-registered with Peer Community in Registered Reports (PCI) (36). Informed consent was obtained from all participants. Unmedicated adults between the ages of 18 and 40 with corrected visual acuity ≥20/35 for each eye (Snellen near vision chart) were recruited from the Greater Toronto Area, including i) individuals with a DSM-5 diagnosis of BDD with primary face concerns and a score of ≥20 on the Yale-Brown Obsessive-Compulsive Scale Modified for BDD (BDD-YBOCS) (37), or ii) individuals with subclinical, defined as a score ≥8 on the Dysmorphic Concern Questionnaire (DCQ) (38) without meeting full DSM-5 criteria for BDD or for any other psychiatric disorder. The subclinical group was included to examine relationships to body dysmorphic symptoms with reduced confounding from comorbid psychiatric disorders, high degrees of ancillary symptoms such as anxiety and depression, and other latent illness effects (39,40). Additional exclusion criteria are detailed in the registered report (36). Modifications and departures from the preregistered protocol are described in the Supplement.

### Assessments

Prior to enrollment, participants completed a telephone screening and a diagnostic evaluation, including the Diagnostic Interview for Anxiety, Mood, and OCD and Related Neuropsychiatric Disorders (DIAMOND) (41), administered by a clinician. The clinician also administered the BDD-YBOCS and the Brown Assessment of Beliefs Scale (BABS) (42) to assess body dysmorphic symptom severity and insight, respectively. Depressive and anxiety symptoms were measured using the Montgomery-Åsberg Depression Rating Scale (MADRS) (43), and the Hamilton Anxiety Rating Scale (HAM-A) (44) and included as covariates in subsequent analyses. Momentary appearance satisfaction, state anxiety, distress, and face concern were also assessed before and after the session (see Supplement).

### Photo Taking and Processing Procedures

Color photographs of participants’ neutral-expression faces were obtained for the fMRI tasks. Four face angles were captured under two lighting conditions each, yielding eight photographs per participant; white balance was standardized across these, and backgrounds were set to black. Photographs were cropped so that the center of the image was between the two eyes (i.e., the bridge of the nose). For control stimuli, each photograph was phase-scrambled using a fast Fourier transform, preserving the original’s color, luminance, and frequency spectrum. The ModV paradigm used the same stimuli overlaid with a translucent crosshair centered on the image. In total, photograph processing yielded 32 images per participant: eight own-face, eight own-face with crosshair, eight phase-scrambled, and eight phase-scrambled with crosshair.

### Face Inversion Effect Task

Global and local processing biases were assessed with the face inversion effect task, a two-alternative forced-choice recognition test consisting of upright and inverted faces (11), administered on a laptop using E-Prime (v2.0; Psychology Software Tools, Inc.). Participants were seated 50 cm from the screen and completed four practice trials before starting the task, which was administered before the TBS session and after the ModV paradigm. The task consisted of four pseudo-randomized blocks, each consisting of upright or inverted faces presented for either a short (500 ms) or a long (5000 ms) duration. Each block contained 28 trials, and each trial consisted of a target face (short or long duration) followed by two selection faces (3000 ms). Participants were instructed to indicate which of the selection faces was the same as the target face as quickly and as accurately as possible. All participants received the same order of pseudo-randomized blocks. Incorrect selection faces were created by morphing each of the individual correct selection faces 50% with another gender-matched face, creating a more difficult identification task (see Feusner, Moller, et al., 2010 for details).

### Initial MRI Procedures

Prior to the TBS procedures, anatomical MRI data were collected using a Siemens 3T *Prisma* scanner with a 32-channel head coil. Specifically, T1-weighted MPRAGE (*tfl3d1; TR/TE: 2300/2.27 ms; flip angle: 8°; 256 x 256 matrix; voxel size: 1 mm^3^; 192 slices) images were collected and used for neuronavigation and registration purposes.

### Theta Burst Stimulation (TBS) Procedures

Neurostimulation was administered using a MagPro R30 (MagVenture, Farum, Denmark) stimulator equipped with a fluid-cooled 70-mm figure-of-eight coil (cool-B70). Resting motor threshold (MT) was determined for both hemispheres with the BrainSight2 (Rogue Research, Montreal, Canada) built-in two-channel electromyography (EMG) device. Readings were taken from the opposite abductor pollicis brevis muscle using pre-gel disposable surface electrodes while stimulation was delivered over the respective motor cortex. Resting MT was defined as the minimum intensity required to evoke a motor evoked potential (MEP) >50 μV in 5 of 10 consecutive trials (45).

T1-weighted images were loaded into the BrainSight2 software and registered to MNI space after manual identification of the anterior and posterior commissures, from which a three-dimensional reconstruction of the participant’s scalp and brain was derived to define stimulation target sites. The BrainSight2 neuronavigation system facilitated MRI-guided coil placement at target foci: left lateral parietal cortex (−38, −38, 46) and right lateral parietal cortex (32, −44, 46) (see Figure 1). Stimulation was delivered at 100% of resting MT. All participants tolerated stimulation for the full duration of both sessions, with no intensity reductions or early terminations. Stimulation parameters for each TBS session consisted of 600 pulses delivered in triplet 50 Hz bursts, repeated at 5 Hz for 2 seconds; for iTBS, each burst was repeated after an 8 s rest, while bursts were applied continuously for cTBS.

**Figure 1.**
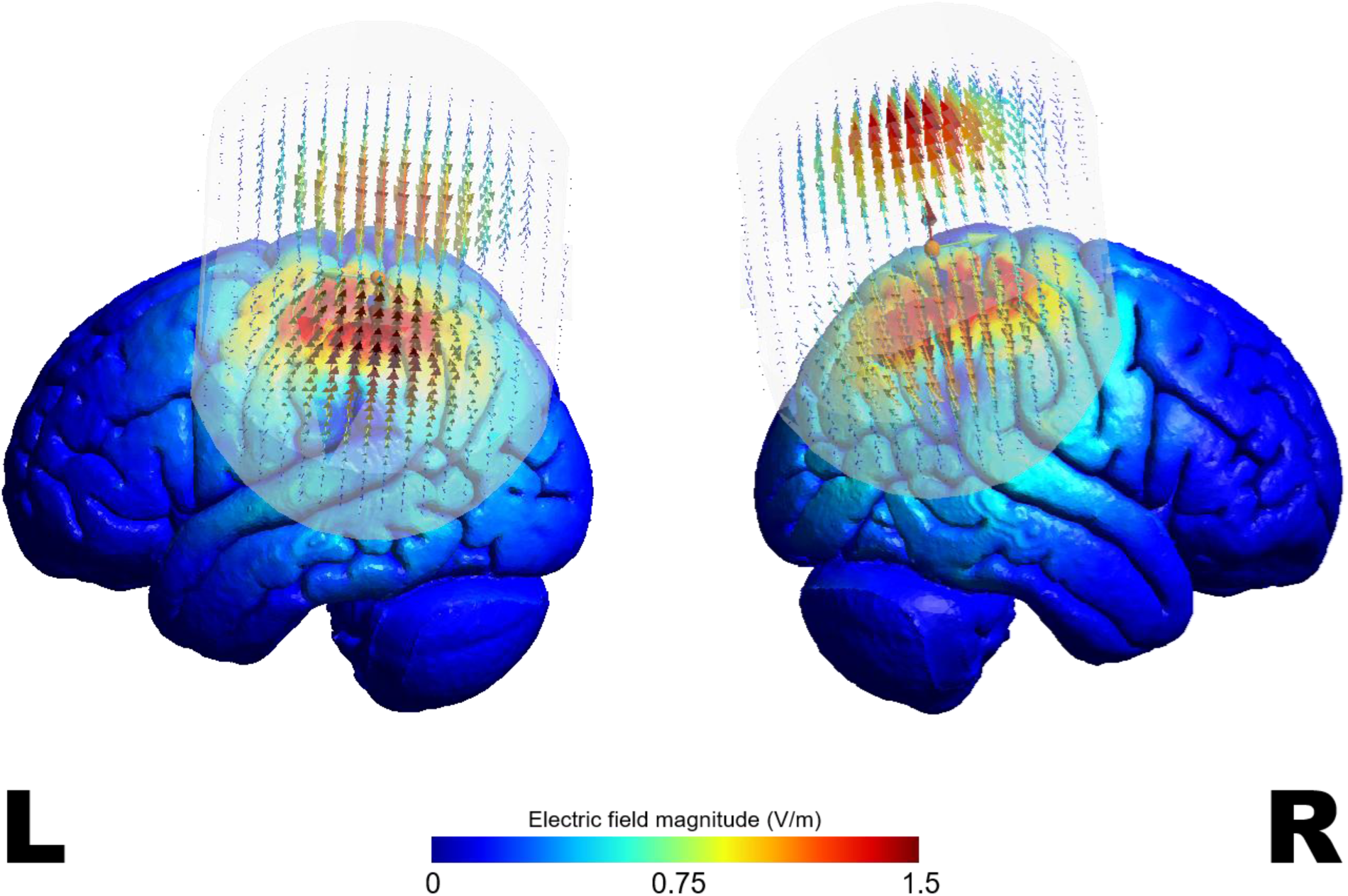
Simulated electric fields for left and right lateral parietal cortex stimulation, determined from meta-analysis of functional brain imaging studies for “dorsal visual stream” (Neurosynth; https://neurosynth.org/), and corresponding to CP3 and CP4 respectively on the EEG 10-10 system. Simulations were generated using SimNIBS v4.6 and projected onto a standard Montreal Neurological Institute (MNI) template brain.

A sham-controlled design using iTBS was initially considered, but active versus sham TBS (e.g. 10% of MT) produce markedly different sensations, and expectations about simulation effects can themselves influence outcomes (46), potentially confounding the design. We instead chose to test the dissociable effects of iTBS and cTBS, which have been shown to increase and decrease functional connectivity, respectively, in specific brain circuits (47,48). The opposite effects from cTBS would provide additional mechanistic proof of concept.

Covariate adaptive randomization determined the order in which participants received iTBS and cTBS, as well as which hemisphere were stimulated during each session. Sessions were separated by a minimum of one day to eliminate carry-over effects. Randomization was counterbalanced within BDD and subclinical BDD groups, with gender balanced as a covariate. Participants, research assistants conducting the tasks, data analysts, and the primary investigator and co-investigator were blinded to condition until data analysis was complete to reduce placebo effects and bias.

### fMRI Acquisition and Procedures

Task-based functional MRI data were acquired with a 64-channel head coil using an HCP multiband sequence (https://www.cmrr.umn.edu/multiband): T2*-weighted echo planar imaging sequence (TR/TE: 1000/30 ms; multi-band accel. factor: 5; flip angle: 60°; 104 x 104 matrix; voxel size: 2 mm^3^; 65 slices). Data acquisition took place a mean of 33.85±17.73 minutes of administration of TBS (the effects of the stimulation may last beyond one hour) (34). fMRI tasks were presented on a 32-inch monitor using custom MATLAB (MathWorks, Inc.) scripts. The visual attention modification paradigm consisted of natural viewing and visual modification tasks (see Figure 2C).

**Figure 2.**
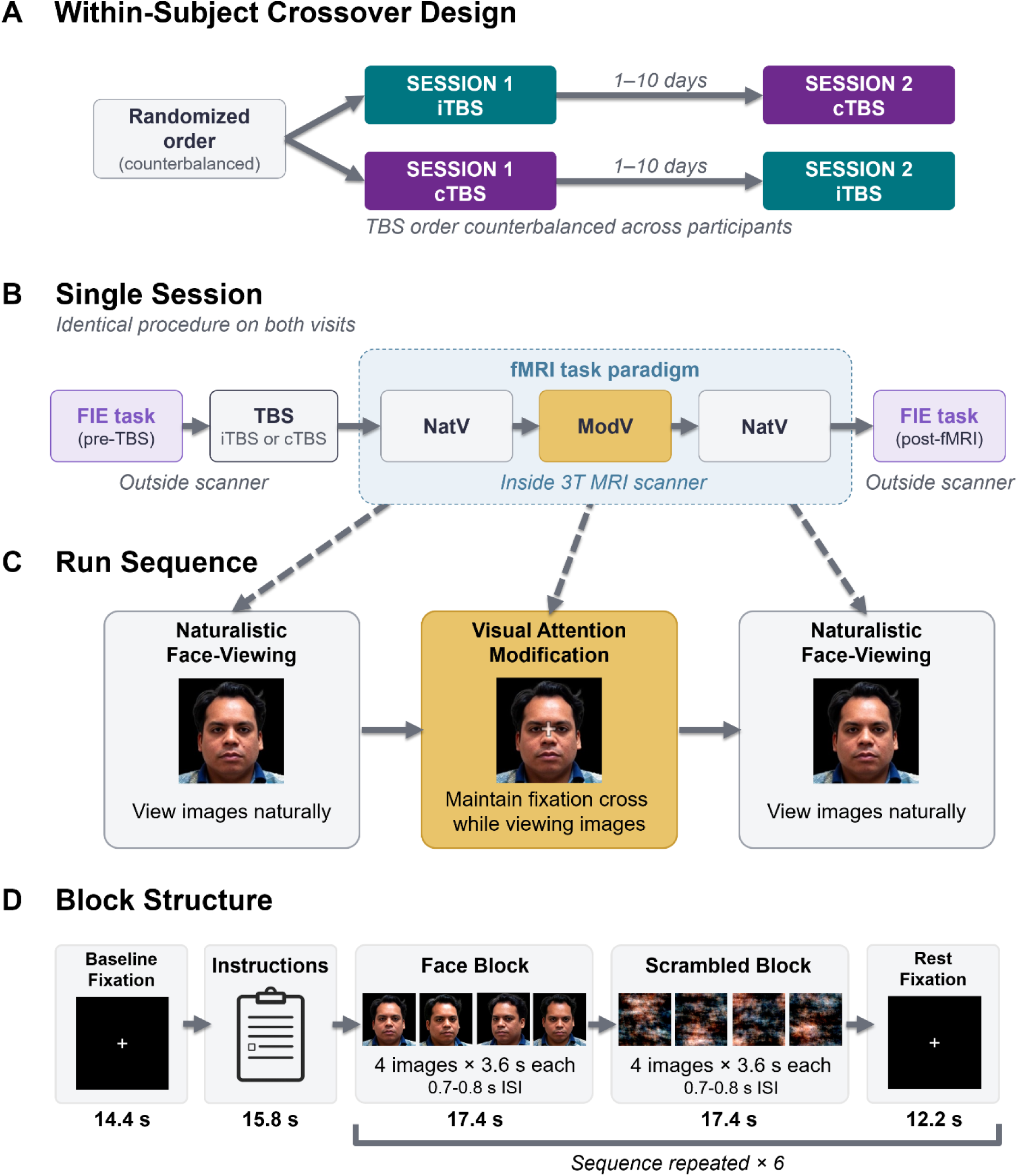
Study design and fMRI task paradigm. **(A)** Participants completed two study visits separated by 1–10 days, receiving intermittent theta burst stimulation (iTBS) during one session and continuous theta burst stimulation (cTBS) during the other in a counterbalanced order. **(B)** During each visit, participants completed the face inversion effect (FIE) task before stimulation, followed by fMRI scanning consisting of an initial Naturalistic Face-Viewing (NatV) run, a Visual Attention Modification (ModV) run, and a second NatV run, after which the FIE task was repeated. **(C)** During the first NatV run, participants were instructed to view photographs of their own face and scrambled control images naturally, as they normally would, and press a button whenever an image appeared to ensure engagement and compliance. During the ModV run, participants viewed the same images while maintaining fixation on a translucent crosshair positioned at the center of each image. Following ModV, the NatV task was repeated and participants were again instructed to view the images naturally. **(D)** A block design was used to present both participant face stimuli and scrambled images for 3.6 s each with 0.7 to 0.8 s between image presentations. Following each block, a fixation cross was presented for 12.2 s. The example participant shown is the author (JPD), who provided consent for the use and publication of these images.

#### fMRI Processing and Connectivity Analysis

Images were processed using *fMRIPrep* v25.1.3 (49). Spatial normalization of the T1-weighted image to standard MNI space was performed through nonlinear registration. Processing of the BOLD timeseries included head-motion estimation, slice time correction, and susceptibility distortion correction using two spin echo field maps of opposite phase encoding directions (see Supplemental Materials for additional fMRIPrep details). Functional data were denoised using probabilistic independent component analysis implemented in FSL MELODIC, followed by automated component classification with ICA-FIX (50) (see Supplemental Materials for details on the custom ICA-FIX classifier). Spatial smoothing was then performed using a Gaussian kernel with a full width at half maximum (FWHM) of 6 mm. The resulting data were used for all subsequent analyses.

The dynamic effective connectivity analysis strategy was based on previous studies (32,51,52). Briefly, blind deconvolution (53) was performed on timeseries extracted from 14 regions of interest (ROIs) (see Figure 3). Using custom MATLAB scripts, dynamic effective connectivity between ROI pairs was computed at each time point using time-varying Granger causality and a dynamic multivariate autoregressive (dMVAR) model, solved in a Kalman-filter framework (54). This approach yielded time-varying model coefficients for each time point in the series. Time points corresponding to face-viewing trials were extracted for higher- and lower-level dorsal and ventral visual stream connections. Positive dynamic effective connectivity values indicate a facilitatory influence, whereby activity in the source region predicts subsequent activity in the target region in the same direction, whereas negative values indicate a suppressive influence. Because the present study focused on feedforward facilitatory signaling, analyses were restricted to positive dynamic effective connectivity values. Connectivity values were subsequently transformed using ordered quantile normalization implemented in the *bestNormalize* R package (55), a rank-based inverse normal transformation mapping the empirical distribution onto a standard normal distribution.

**Figure 3.**
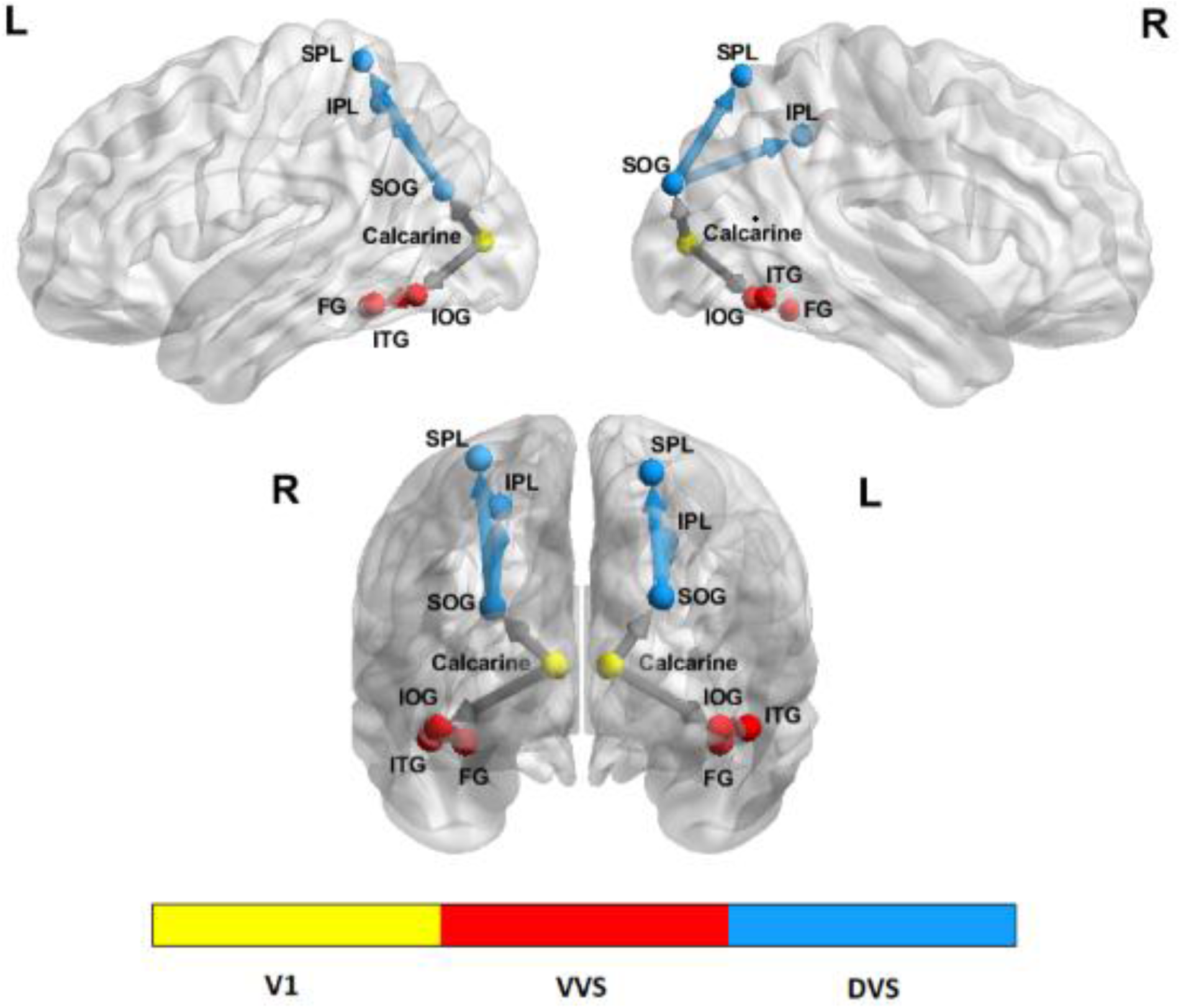
Fourteen regions of interest (ROIs) were included in the connectivity analyses. ROIs were derived from a Neurosynth functional meta-analysis in the dorsal and ventral visual streams using maps generated through association tests acquired using search queries including "primary visual," "ventral visual," "visual stream," and "dorsal visual”. The ROI set comprised V1 [bilateral calcarine], six ventral visual stream (VVS) ROIs [bilateral inferior occipital gyrus (IOG), fusiform gyrus (FG), and inferior temporal gyrus (ITG)], and six dorsal visual stream (DVS) ROIs [bilateral superior occipital gyrus (SOG), inferior parietal lobule (IPL), and superior parietal lobule (SPL)]. Twelve intra-hemispheric connections were examined and grouped into four categories: lower DVS [calcarine to SOG], higher DVS [SOG to IPL and SPL], lower VVS [calcarine to IOG], and higher VVS [IOG to FG and ITG]. All ROIs were defined as 5 mm radius spheres. MNI coordinates are provided in Table S1.

### Statistical Analysis

To examine changes in dynamic effective connectivity during naturalistic own-face viewing, separate linear mixed models (LMM) were used for each TBS condition, with task condition (naturalistic own-face viewing before or after ModV), visual stream level (dorsal or ventral; lower or higher), and their interactions as fixed factors, participant as a random factor, and head motion (DVARS) as a covariate of non-interest. Pairwise comparisons were performed for significant interaction effects.

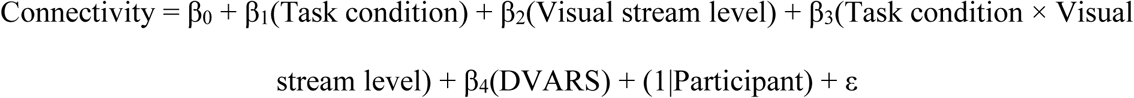

For each LMM, we assessed potential sources of interindividual variability, including age, gender, time of day, time elapsed between stimulation and scan, TBS order, resting MT, and depression and anxiety measures; covariates were retained in the final model only if statistically significant (*p* < .05) to maintain model parsimony and interpretability.

To examine the effects of stimulation on global visual processing biases, face inversion task response times faster than 300 ms were excluded, as such responses are unlikely to reflect genuine stimulus-driven processing and more likely represent anticipatory or accidental responses. The remaining response times were log-transformed to reduce positive skew and better satisfy the assumptions of parametric statistical analyses. Face inversion effect scores were then calculated from the log-transformed response times. Paired t-tests were performed on residualized face inversion effect scores to compare performance before and after iTBS and cTBS, controlling for task order and the interval between stimulation and task completion.

## Results

### Sample Characteristics

Twenty-nine participants with BDD and 10 with subclinical BDD symptoms underwent MRI scanning. One participant with BDD was excluded due to substantial signal dropout across all scans. One additional participant with BDD completed only an iTBS session, and another had a truncated visual attention modification scan; available data from these participants were retained where applicable. The final analytic sample included 38 participants (Table 1).

**Table 1.**
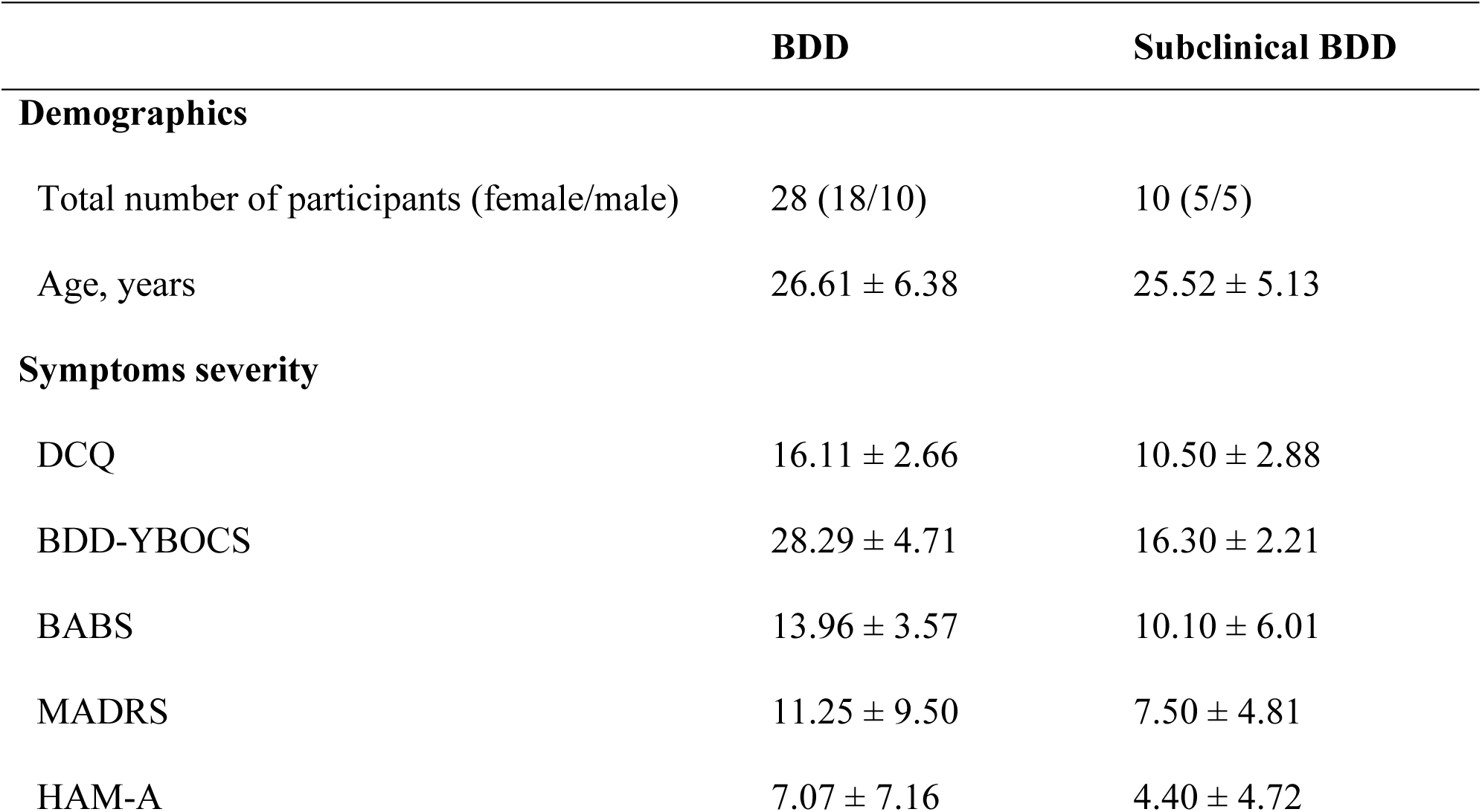
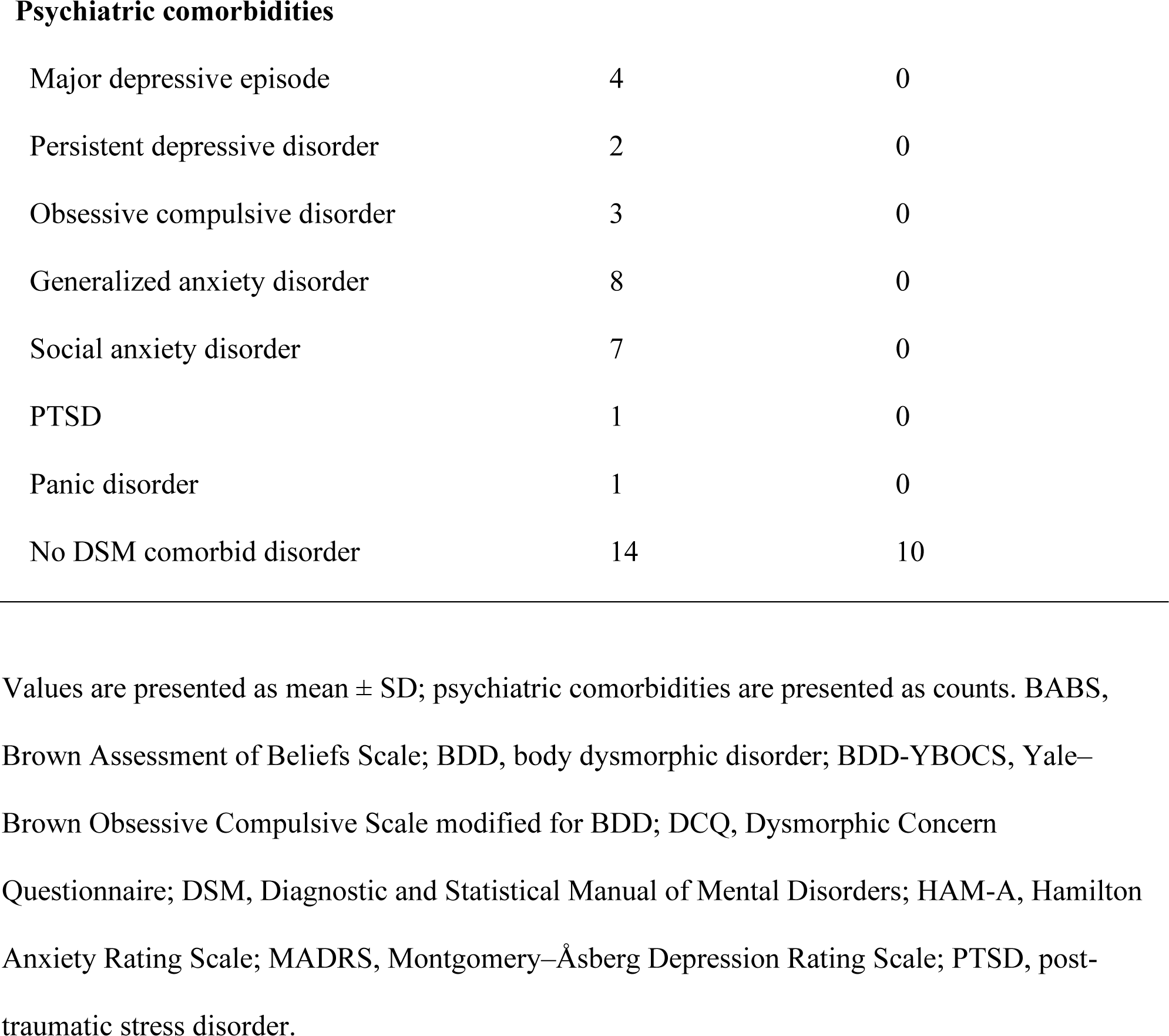
Sample Characteristics.

|  | BDD | Subclinical BDD |
| --- | --- | --- |
| **Demographics** |  |  |
| Total number of participants (female/male) | 28 (18/10) | 10 (5/5) |
| Age, years | 26.61 ± 6.38 | 25.52 ± 5.13 |
| **Symptoms severity** |  |  |
| DCQ | 16.11 ± 2.66 | 10.50 ± 2.88 |
| BDD-YBOCS | 28.29 ± 4.71 | 16.30 ± 2.21 |
| BABS | 13.96 ± 3.57 | 10.10 ± 6.01 |
| MADRS | 11.25 ± 9.50 | 7.50 ± 4.81 |
| HAM-A | 7.07 ± 7.16 | 4.40 ± 4.72 |
| Psychiatric comorbidities |  |  |
| Major depressive episode | 4 | 0 |
| Persistent depressive disorder | 2 | 0 |
| Obsessive compulsive disorder | 3 | 0 |
| Generalized anxiety disorder | 8 | 0 |
| Social anxiety disorder | 7 | 0 |
| PTSD | 1 | 0 |
| Panic disorder | 1 | 0 |
| No DSM comorbid disorder | 14 | 10 |

Values are presented as mean ± SD; psychiatric comorbidities are presented as counts. BABS, Brown Assessment of Beliefs Scale; BDD, body dysmorphic disorder; BDD-YBOCS, Yale–Brown Obsessive Compulsive Scale modified for BDD; DCQ, Dysmorphic Concern Questionnaire; DSM, Diagnostic and Statistical Manual of Mental Disorders; HAM-A, Hamilton Anxiety Rating Scale; MADRS, Montgomery–Åsberg Depression Rating Scale; PTSD, post-traumatic stress disorder.

### iTBS-related Dynamic Effective Connectivity Changes

Following iTBS, dynamic effective connectivity showed selective modulation across visual pathways (Figure 4A). During naturalistic own-face viewing post-visual attention modification compared with pre-visual attention modification, connectivity increased in the higher level dorsal visual stream (DVS higher: β = .079, *p* = .008) and in the lower-level ventral visual stream (VVS lower: β = .110, *p* < .001). No significant changes were observed for DVS lower (β = −.027, *p* = .352) or VVS higher pathways (β = −.025, *p* = .328). In addition, greater head motion (DVARS) was associated with higher connectivity estimates (β = .058, *p* = .003).

**Figure 4.**
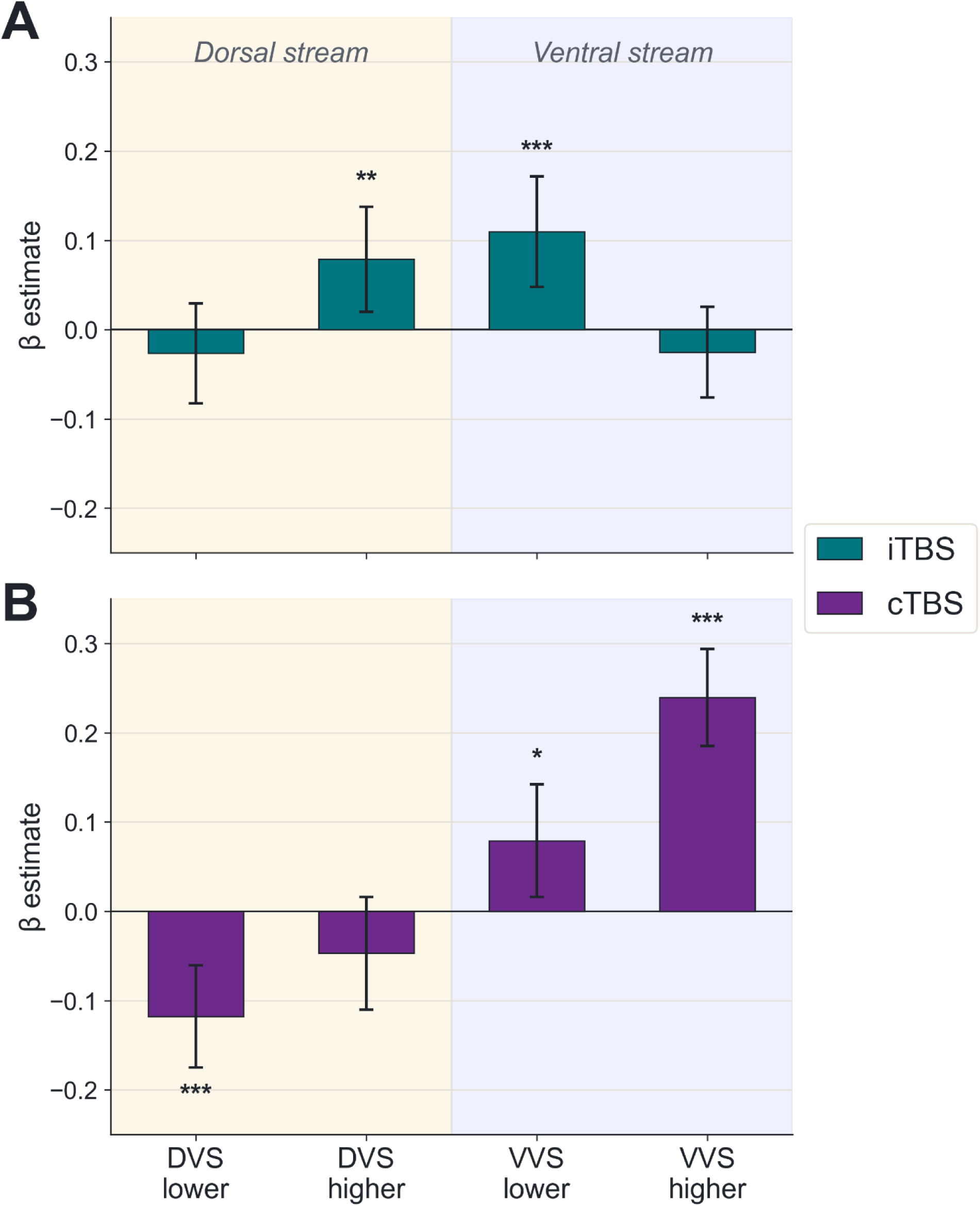
Changes in dynamic effective connectivity patterns across runs in dorsal and ventral visual stream pathways following theta burst stimulation prior to the own-face viewing attention modulation task. **(A)** iTBS-related connectivity changes in dorsal and ventral visual streams. **(B)** cTBS-related connectivity changes in dorsal and ventral visual streams. Bars represent fixed-effects β estimates for the change in connectivity from before to after attention modification during naturalistic own-face viewing (post-ModV − pre-ModV; i.e., the second versus first naturalistic viewing run), estimated from separate linear mixed-effects models for each stimulation condition. Positive values indicate increased connectivity following attention modification, whereas negative values indicate decreased connectivity. Error bars indicate 95% confidence intervals. Statistically significant effects are denoted by \**p* < .05, \*\**p* < .01, \*\*\**p* < .001. cTBS, continuous theta burst stimulation; DEC, dynamic effective connectivity; DVS, dorsal visual stream; iTBS, intermittent theta burst stimulation; VVS, ventral visual stream.

### cTBS-related Dynamic Effective Connectivity Changes

Following cTBS, dynamic effective connectivity showed distinct modulation across visual pathways (Figure 4B). During naturalistic own-face viewing post-visual attention modification compared with pre-visual attention modification, dynamic effective connectivity decreased in the dorsal stream at the lower level (DVS lower: β = −.118, *p* < .001), while the ventral stream showed significant increases at the higher level (VVS higher: β = .239, *p* < .001) and at the lower level (VVS lower: β = .079, *p* = .014). The change in DVS higher was not significant (β = −.047, *p* = .146) (Figure 4B). Among the retained covariates, higher depression severity (MADRS) was associated with lower connectivity estimates (β = −.063, *p* = .005), whereas higher resting MT was associated with greater connectivity (β = .047, *p* = .031). Head motion (DVARS) was not significantly associated with connectivity (β = .016, *p* = .329).

### Face Inversion Effect Changes Following TBS

Paired-samples comparisons evaluated changes in face inversion effect response times before versus after TBS with visual modification. Following iTBS, there was a significant decrease in the face inversion effect for short-duration trials (*t*(38) = −2.44, *p* = .019, mean Δ = −.058), with a lesser magnitude, nonsignificant effect for long-duration trials (*t*(38) = −1.37, *p* = .180, mean Δ = −.039). Following cTBS, there was no significant change for short-duration trials (*t*(37) = −1.48, *p* = .146, mean Δ = −.040), but a significant decrease for long-duration trials (*t*(37) = −2.14, *p* = .039, mean Δ = −.053).

As a post hoc exploratory analysis, we assessed whether stimulation-related changes differed from expected face inversion task practice effects from repeated task exposure. We compared iTBS-first (*n* = 20) and cTBS-first (*n* = 19) subgroups against a separate no-stimulation dataset collected using the same task at two timepoints in individuals with BDD and subclinical BDD (*n* = 37), enrolled under the same inclusion/exclusion criteria and similar demographic characteristics (Table S6). The largest numerical divergence from the no-stimulation condition was observed for short-duration trials following iTBS and long-duration trials following cTBS. For short-duration trials, the no-stimulation group demonstrated a mean pre- to post-session reduction in the face inversion effect of Δ = −.028, whereas the iTBS-first subgroup demonstrated a smaller reduction (Δ = −.009). For long-duration trials, the cTBS-first subgroup demonstrated a larger reduction (Δ = −.084) relative to the no-stimulation group (Δ = −.015). However, the stimulation × session interactions were not statistically significant for either the iTBS short-duration comparison (*p* = .652) or the cTBS long-duration comparison (*p* = .164). Together, these exploratory findings suggest an increase in the face inversion effect at the short duration following iTBS relative to practice effects (repeated face inversion task with no stimulation in-between), and a decrease in the face inversion effect at the long duration following cTBS relative to practice effects (full statistical results are provided in the Supplementary Materials).

## Discussion

This is the first study to investigate the differential effects of a single session of iTBS and cTBS, combined with visual attention modification during self-face viewing, on dynamic effective connectivity within dorsal and ventral visual streams and on global visual processing biases in individuals with BDD symptoms. iTBS selectively enhanced higher-level dorsal and lower-level ventral stream connectivity, whereas cTBS reduced lower-level dorsal connectivity alongside increased lower and higher-level ventral connectivity. Behaviorally, a decreased face inversion effect was observed after iTBS for short face-presentation durations and after cTBS for longer durations. However, accounting for practice effects, iTBS produced a small relative enhancement of the face inversion effect at short durations and cTBS a small relative reduction at long durations.

The central pattern suggests that stimulation alters the relative engagement of dorsal (global, spatial integration) and ventral (local, feature-based) processing streams (56) in a level-specific manner. These TBS-related changes therefore do not map cleanly onto dichotomous increases or decreases in “global” versus “local” connections, but instead reflect selective modulation of hierarchical integration within each stream. Increased higher-level dorsal connectivity following iTBS may index enhanced global integration of spatial or configural information (57), whereas increased ventral connectivity, particularly under cTBS, may reflect greater reliance on lower-level feature processing, as proposed in models of visual processing abnormalities in BDD (58).

Although these exploratory behavioral comparisons were not statistically significant, the directional pattern may nevertheless be mechanistically informative. Short-duration face inversion effect is typically interpreted as reflecting rapid, automatic configural processing, whereas longer durations allow for more strategic or feature-based processing (11,28). Within this framework, iTBS may have relatively increased the balance of global to local processing, whereas cTBS may have promoted later-stage detail-oriented processing. Clinically, iTBS’s effect on brief, automatic viewing could translate to more holistic incidental self-perception, such as a quick glance in a mirror, whereas cTBS’s effect at longer durations may parallel the sustained, feature-focused scrutiny of mirror checking and other prolonged self-inspection common in BDD. Importantly, these effects followed only a single session, and larger or more consistent effects may require repeated sessions (59).

Visual attention modification alone reduces eye scanning behavior and increase connectivity between early visual cortex and dorsal stream regions during subsequent naturalistic self-face viewing, suggesting enhanced global processing (29). A prior TBS pilot study in BDD demonstrated that excitatory stimulation can increase dorsal stream connectivity during both natural face viewing (32) and rapid, short-duration face viewing (52). The present results extend this evidence by showing that combining the two produces pathway-specific connectivity changes aligning with possible shifts in perceptual processing. One plausible interpretation is that attention modification establishes a network state that is then amplified or reconfigured by pre-application of TBS (60,61). Under iTBS, this may facilitate propagation of information through higher-level dorsal pathways, enhancing integrative processing; under cTBS, the same state may be redirected toward ventral pathways, reinforcing feature-based processing. Given that BDD is characterized by relatively reduced dorsal and enhanced ventral stream engagement (20,21), selectively strengthening dorsal, integrative processing with iTBS suggests a candidate mechanism for correcting this imbalance that combined neuromodulation and attentional retraining could exploit therapeutically.

These findings also support a mechanistic rationale for testing combined neuromodulation with perceptual retraining as a potential intervention in BDD, although the magnitude of effects observed here suggests that repeated sessions would be needed for clinically meaningful change. Importantly, although current evidence-based treatments for BDD, including serotonin reuptake inhibitors and cognitive behavioral therapy, can improve symptoms (62), they have not been definitively shown to directly modify the underlying perceptual distortions of appearance (63), raising the possibility that neuromodulation-based interventions may target a mechanistic domain that remains insufficiently addressed.

Several limitations constrain interpretation. First, there was no condition isolating TBS effects in the absence of visual attention modification, making it difficult to disentangle independent versus interactive effects. Second, although the intent of this experiment was to establish early mechanistic plausibility, the use of a single TBS session limits conclusions about durability and clinical relevance. Finally, estimation of practice effects were derived from a comparison study with a different inter-task interval than the current protocol. However, because practice effects may be greater over shorter intervals, this approach may have underestimated practice-related improvements in the present study, and thus underestimated the relative increase in global processing from iTBS plus attention modification.

In summary, iTBS and cTBS applied to lateral parietal cortex, when combined with visual attention modification, produced distinct, pathway-specific alterations in dynamic effective connectivity within visual processing networks, accompanied by temporally specific shifts in perceptual processing, suggesting that neuromodulation could influence the balance of hierarchical information flow underlying visual perception in BDD. The findings provide proof-of-concept for targeting visual network dynamics through combined neuromodulatory and perceptual interventions, with potential implications for developing mechanistically informed treatments for disorders of body image.

## Acknowledgments

Funding for this study is provided by the National Institute of Mental Health (NIMH) grant number: R21MH128815 (Feusner). Joel P. Diaz-Fong was supported in part by the International OCD Foundation 2025 Michael A. Jenike Young Investigator Award. The authors thank Arla Dakli, Matthew Kuo, Jacy Wang, and Daren Liang for their contributions to project administration and investigation, including participant coordination and data collection. This research was enabled in part by support provided by Compute Ontario (<u>computeontario.ca</u>) and the Digital Research Alliance of Canada (<u>alliancecan.ca</u>).

## Disclosures

### CRediT Authorship Contribution Statement

**Joel P. Diaz-Fong:** Formal analysis, Investigation, Data Curation, Visualization, Writing – original draft, Writing – review & editing. **Hayden J. Peel:** Formal analysis, Writing – review & editing. **Kaixi Zhang:** Formal analysis, Writing – review & editing. **Jessica Qian:** Investigation, Data Curation, Visualization, Writing – review & editing. **Madison Lewis:** Investigation, Visualization, Writing – review & editing. **Wan-Wa Wong:** Conceptualization, Methodology, Writing – review & editing. **Andrew F. Leuchter:** Conceptualization, Methodology, Writing – review & editing. **Reza Tadayonnejad:** Conceptualization, Methodology. **Daphne Voineskos:** Investigation, Resources, Supervision, Writing – review & editing. **Gerasimos Konstantinou:** Investigation, Writing – review & editing. **Eileen Lam:** Investigation, Project administration, Writing – review & editing. **Daniel M. Blumberger:** Resources, Writing – review & editing. **Jamie D. Feusner:** Conceptualization, Funding acquisition, Investigation, Methodology, Supervision, Writing – original draft, Writing – review & editing.

### Data Availability Statement

The data supporting the findings of this study are available through the National Institute of Mental Health (NIMH) Data Archive (https://nda.nih.gov/; Collection ID: 4393), subject to controlled access in accordance with NIMH Data Archive policies.

