## Supplemental Materials for "Effects of theta burst stimulation on neural connectivity and visual perception following attention modification of own-face viewing in body dysmorphic disorder"

[1]Brain Health Imaging Centre, Centre for Addiction and Mental Health, Toronto, Ontario, Canada; [2]Institute of Medical Science, Temerty Faculty of Medicine, University of Toronto, Toronto, Ontario, Canada; [3]Data Sciences Institute, University of Toronto, Toronto, Ontario, Canada; [4]Department of Psychiatry and Biobehavioral Science, David Geffen School of Medicine at University of California Los Angeles, Los Angeles, California, United States; [5]TMS Clinical and Research Service, Neuromodulation Division, Semel Institute for Neuroscience and Human Behavior at University of California Los Angeles, Los Angeles, California, United States; [6]Department of Psychology, University of Toronto Scarborough, Toronto, Ontario, Canada; [7]Hong Kong Center for Neurodegenerative Diseases, Hong Kong Science Park, Hong Kong, China; [8]Temerty Centre for Therapeutic Brain Intervention, Campbell Family Research Institute, Centre for Addiction and Mental Health, Toronto, Ontario, Canada; [9]Poul Hansen Family Centre for Depression, Centre for Mental Health, University Health Network, Toronto, Ontario, Canada; [10]Department of Psychiatry, Temerty Faculty of Medicine, University of Toronto, Toronto, Ontario, Canada; [11]Department of Women's and Children's Health, Karolinska Institutet, Stockholm, Sweden

**Author Note**

Joel P. Diaz-Fong, https://orcid.org/0000-0002-2108-6830
Hayden J. Peel, https://orcid.org/0000-0001-6078-2970
Kaixi Zhang, https://orcid.org/0000-0003-2073-7264
Jessica Qian, https://orcid.org/0009-0008-6000-312X
Madison Lewis, https://orcid.org/0009-0007-1429-0845
Gerasimos Konstantinou, https://orcid.org/0000-0002-0303-1633
Daniel M. Blumberger, https://orcid.org/0000-0002-8422-5818
Jamie D. Feusner, https://orcid.org/0000-0002-0391-345X

**Correspondence**

Jamie D. Feusner, 250 College St. #645, Toronto, Ontario M5T 1R8, Canada.


**Contents:**

### Supplementary Methods and Results

1. Protocol Modifications and Departures from Preregistration
2. Details on fMRIprep preprocessing
3. ICA-FIX Classifier Training and Denoising
4. Linear Mixed-Effects Model Results for Dynamic Effective Connectivity Following iTBS and cTBS
5. Exploratory Post Hoc Analyses of the Face Inversion Effect
6. Self-Reported Mood, Anxiety, Distress, and Appearance Concern

### Supplementary Tables

Table S1. MNI coordinates for regions of interest included in the connectivity analyses
Table S2. Linear mixed-effects model of dynamic effective connectivity following iTBS
Table S3. Linear mixed-effects model of dynamic effective connectivity following cTBS
Table S4. Pairwise comparisons of dynamic effective connectivity between runs within each visual stream level following iTBS
Table S5. Pairwise comparisons of dynamic effective connectivity between runs within each visual stream level following cTBS
Table S6. Sample characteristics of the no-stimulation face inversion effect dataset
Table S7. Body image state and anxiety scores before and after stimulation
Table S8. Distress and face concern ratings during experimental sessions
Table S9. Mixed-effects model results for distress and face concern ratings

### Supplementary Figures

Figure S1. iTBS versus no-stimulation changes in the face inversion effect
Figure S2. cTBS versus no-stimulation changes in the face inversion effect

### Supplementary References

1. Protocol Modifications and Departures from Preregistration

Several modifications to the preregistered protocol were implemented during the course of the study. First, the preregistered recruitment target of 20 individuals with DSM-5 body dysmorphic disorder (BDD) and 20 individuals with subclinical BDD symptoms was not achieved. Recruitment of individuals with subclinical BDD symptoms proved more difficult than anticipated within the available funding period, resulting in a smaller subclinical BDD sample than originally planned. Second, instead of the Depression Anxiety and Stress Scale (DASS-21) (1) as originally planned, depressive and anxiety symptom severity were assessed using the Montgomery-Åsberg Depression Rating Scale (MADRS) and Hamilton Anxiety Rating Scale (HAM-A). Third, preprocessing procedures differed from the preregistered pipeline. Although ICA-AROMA was originally planned for denoising, functional MRI data were instead denoised using ICA-FIX. This decision was made because the multiband acquisition introduced artifacts that were not adequately captured by ICA-AROMA, whereas ICA-FIX provided improved classification and removal of structured noise components in the multiband data. Finally, although the preregistration indicated that participants would be asked whether they believed they had received intermittent theta burst stimulation (iTBS) or continuous theta burst stimulation (cTBS) to assess the effectiveness of blinding, this information was inadvertently not collected.

2. Details on fMRIprep Preprocessing

Results included in this manuscript come from preprocessing performed using *fMRIPrep* 25.1.3 (2), which is based on *Nipype* 1.10.0 (3).

*Preprocessing of $B_0$ inhomogeneity mappings.* A total of 1 fieldmaps were found available within the input BIDS structure for each subject. A $B_0$-nonuniformity map (or *fieldmap*) was estimated based on two (or more) echo-planar imaging (EPI) references with topup (4).

*Anatomical data preprocessing.* A total of 1 T1-weighted (T1w) images were found within the input BIDS dataset. The T1w image was corrected for intensity non-uniformity (INU) with N4BiasFieldCorrection (5), distributed with ANTs 2.6.2 (6), and used as T1w-reference throughout the workflow. The T1w-reference was then skull-stripped with a *Nipype* implementation of the antsBrainExtraction.sh workflow (from ANTs), using OASIS30ANTs as target template. Brain tissue segmentation of cerebrospinal fluid (CSF), white-matter (WM) and gray-matter (GM) was performed on the brain-extracted T1w using fast (FSL (version unknown) (7). Volume-based spatial normalization to two standard spaces (MNI152NLin6Asym, MNI152NLin2009cAsym) was performed through nonlinear registration with antsRegistration (ANTs 2.6.2), using brain-extracted versions of both T1w reference and the T1w template. The following templates were selected for spatial normalization and accessed with *TemplateFlow* (24.2.2) (8): *FSL's MNI ICBM 152 non-linear 6th Generation Asymmetric Average Brain Stereotaxic Registration Model* (9) [TemplateFlow ID: MNI152NLin6Asym], *ICBM 152 Nonlinear Asymmetrical template version 2009c* (10) [TemplateFlow ID: MNI152NLin2009cAsym].

*Functional data preprocessing*. For each of the 8 BOLD runs found per subject (across all tasks and sessions), the following preprocessing was performed. First, a reference volume was generated, using a custom methodology of *fMRIPrep*, for use in head motion correction. Head-motion parameters with respect to the BOLD reference (transformation matrices, and six corresponding rotation and translation parameters) are estimated before any spatiotemporal filtering using mcflirt (FSL) (11). The estimated *fieldmap* was then aligned with rigid-registration to the target EPI (echo-planar imaging) reference run. The field coefficients were mapped on to the reference EPI using the transform. The BOLD reference was then co-registered to the T1w reference using mri_coreg (FreeSurfer) followed by flirt (FSL) (12) with the boundary-based registration (13) cost-function. Co-registration was configured with six degrees of freedom. Several confounding time-series were calculated based on the *preprocessed BOLD*: framewise displacement (FD), DVARS and three region-wise global signals. FD was computed using two formulations following Power (absolute sum of relative motions) (14) and Jenkinson (relative root mean square displacement between affines) (11). FD and DVARS are calculated for each functional run, both using their implementations in *Nipype* (14). The three global signals are extracted within the CSF, the WM, and the whole-brain masks. Additionally, a set of physiological regressors were extracted to allow for component-based noise correction (*CompCor*) (15). Principal components are estimated after high-pass filtering the *preprocessed BOLD* time-series (using a discrete cosine filter with 128s cut-off) for the two *CompCor* variants: temporal (tCompCor) and anatomical (aCompCor). tCompCor components are then calculated from the top 2% variable voxels within the brain mask. For aCompCor, three probabilistic masks (CSF, WM and combined CSF+WM) are generated in anatomical space. The implementation differs from that of Behzadi et al. (15) in that instead of eroding the masks by 2 pixels on BOLD space, a mask of pixels that likely contain a volume fraction of GM is subtracted from the aCompCor masks. This mask is obtained by thresholding the corresponding partial volume map at 0.05, and it ensures components are not extracted from voxels containing a minimal fraction of GM. Finally, these masks are resampled into BOLD space and binarized by thresholding at 0.99 (as in the original implementation). Components are also calculated separately within the WM and CSF masks. For each CompCor decomposition, the *k* components with the largest singular values are retained, such that the retained components' time series are sufficient to explain 50 percent of variance across the nuisance mask (CSF, WM, combined, or temporal). The remaining components are dropped from consideration. The head-motion estimates calculated in the correction step were also placed within the corresponding confounds file. The confound time series derived from head motion estimates and global signals were expanded with the inclusion of temporal derivatives and quadratic terms for each (16). Frames that exceeded a threshold of 0.5 mm FD or 1.5 standardized DVARS were annotated as motion outliers. Additional nuisance timeseries are calculated by means of principal components analysis of the signal found within a thin band (*crown*) of voxels around the edge of the brain, as proposed by (17). All resamplings can be performed with *a single interpolation step* by composing all the pertinent transformations (i.e. head-motion transform matrices, susceptibility distortion correction when available, and co-registrations to anatomical and output spaces). Gridded (volumetric) resamplings were performed using nitransforms, configured with cubic B-spline interpolation. Many internal operations of *fMRIPrep* use *Nilearn* 0.11.1 (18), mostly within the functional processing workflow.

3. ICA-FIX Classifier Training and Denoising

ICA was performed on unsmoothed data with a high-pass filter of 100 s, and dimensionality was estimated automatically. A subset of 20 runs across participants was manually labeled into signal and noise components based on established spatial and temporal criteria for ICA component classification (e.g., motion-related edge artifacts, high-frequency noise, and non-gray matter localization) (19). These labels were used to train a study-specific FIX classifier using the default feature set. Classifier performance was assessed using leave-one-out cross-validation. At a classification threshold of 20, the classifier achieved a weighted accuracy of 99%, with a true positive rate of 100% and a true negative rate of 97%. The trained classifier was then applied to all runs, and noise components were removed using non-aggressive regression. In addition, six rigid-body motion parameters (three translations and three rotations) were included as nuisance regressors and regressed from the time series.

***Supplementary Table S1.*** **MNI coordinates for the regions of interest included in the connectivity analyses**

| **Region of Interest (ROI)** | | **MNI Coordinates** | |
|---|---|---|---|
| | **X** | **Y** | **Z** |
| L Calcarine | −8 | −86 | 6 |
| L Inferior Occipital Gyrus (IOG) | −42 | −64 | −12 |
| L Fusiform Gyrus (FG) | −34 | −48 | −16 |
| L Inferior Temporal Gyrus (ITG) | −44 | −50 | −15 |
| L Superior Occipital Gyrus (SOG) | −26 | −73 | 23 |
| L Inferior Parietal Lobule (IPL) | −24 | −52 | 52 |
| L Superior Parietal Lobule (SPL) | −30 | −46 | 66 |
| R Calcarine | 8 | −86 | 6 |
| R Inferior Occipital Gyrus (IOG) | 40 | −64 | −12 |
| R Fusiform Gyrus (FG) | 40 | −52 | −16 |
| R Inferior Temporal Gyrus (ITG) | 48 | −60 | −12 |
| R Superior Occipital Gyrus (SOG) | 23 | −91 | 26 |
| R Inferior Parietal Lobule (IPL) | 24 | −48 | 42 |
| R Superior Parietal Lobule (SPL) | 20 | −68 | 62 |

L, left hemisphere; MNI, Montreal Neurological Institute; R, right hemisphere

4. Linear Mixed-Effects Model Results for Dynamic Effective Connectivity Following iTBS and cTBS

*Dynamic Effective Connectivity Analysis*. Dynamic effective connectivity during own-face viewing was analyzed using separate linear mixed-effects models for iTBS and cTBS sessions. Connectivity values were restricted to positive estimates, averaged across hemispheres, and normalized with an ordered quantile (ORQ) transformation. The dependent variable was normalized dynamic effective connectivity. Fixed effects included visual stream level (four levels reflecting the lower and higher levels of the dorsal visual stream and ventral visual stream) and its interaction with run (Natural Face-Viewing 1, Visual Attention Modification, and Natural Face-Viewing 2), with Natural Face-Viewing 1 as the reference run. A by-participant random intercept was included. Models were estimated by restricted maximum likelihood (REML), and denominator degrees of freedom for fixed effects were approximated using the Satterthwaite method. Candidate covariates of non-interest (age, gender, time of day, time elapsed between stimulation and scan, TBS order, resting motor threshold, and depression and anxiety measures) were evaluated and retained only if they reached statistical significance ($p < .05$). DVARS was included as a covariate in all models to account for head motion. Consequently, the final iTBS model included DVARS, whereas the final cTBS model included resting motor threshold, depression severity (MADRS), and DVARS. Full fixed-effects estimates for the iTBS and cTBS models are reported in Supplementary Tables S2 and S3, respectively.

Estimated marginal means were computed from each fitted model, and pairwise comparisons between runs were performed within each visual stream level for significant interaction effects. Because the number of observations exceeded the threshold for exact degree-of-freedom estimation, these contrasts used asymptotic ($z$) tests, and $p$ values were adjusted for multiple comparisons within each visual stream level using the Holm method. Holm-adjusted pairwise comparisons of estimated marginal means are reported in Supplementary Tables S4 and S5.

***Supplementary Table S2.* Linear mixed-effects model of dynamic effective connectivity following iTBS**

| **Fixed effect** | **Estimate** | **SE** | **df** | ***t*** | ***p*** |
|---|---|---|---|---|---|
| DVS higher, NatV1 (reference) | −0.369 | 0.034 | 79.3 | −10.945 | **<.001** |
| DVS lower, NatV1 (reference) | 0.142 | 0.033 | 70.9 | 4.344 | **<.001** |
| VVS higher, NatV1 (reference) | 0.143 | 0.032 | 63.4 | 4.491 | **<.001** |
| VVS lower, NatV1 (reference) | −0.114 | 0.035 | 87.7 | −3.286 | **.001** |
| DVARS | 0.058 | 0.019 | 138.4 | 3.070 | **.003** |
| DVS higher, ModV vs. NatV1 | 0.053 | 0.030 | 27830 | 1.749 | .080 |
| DVS lower, ModV vs. NatV1 | 0.002 | 0.028 | 27830 | 0.077 | .939 |
| VVS higher, ModV vs. NatV1 | 0.011 | 0.026 | 27830 | 0.432 | .665 |

| | | | | | |
|---|---|---|---|---|---|
| VVS lower, ModV vs. NatV1 | 0.086 | 0.031 | 27830 | 2.741 | **.006** |
| DVS higher, NatV2 vs. NatV1 | 0.079 | 0.030 | 27830 | 2.634 | **.008** |
| DVS lower, NatV2 vs. NatV1 | −0.027 | 0.029 | 27820 | −0.931 | .352 |
| VVS higher, NatV2 vs. NatV1 | −0.025 | 0.026 | 27770 | −0.977 | .328 |
| VVS lower, NatV2 vs. NatV1 | 0.110 | 0.031 | 27810 | 3.487 | **<.001** |

Bold *p* values indicate statistical significance ($p < .05$). df, Satterthwaite degrees of freedom; DVARS, head motion; DVS, dorsal visual stream; ModV, Visual Attention Modification run; NatV1, first Natural Face-Viewing run; NatV2, second Natural Face-Viewing run; VVS, ventral visual stream.

***Supplementary Table S3.*** **Linear mixed-effects model of dynamic effective connectivity following cTBS**

| **Fixed effect** | **Estimate** | **SE** | **df** | ***t*** | ***p*** |
|---|---|---|---|---|---|
| DVS higher, NatV1 (reference) | −0.245 | 0.030 | 138.0 | −8.156 | **<.001** |
| DVS lower, NatV1 (reference) | 0.173 | 0.028 | 100.3 | 6.226 | **<.001** |
| VVS higher, NatV1 (reference) | −0.061 | 0.028 | 104.6 | −2.160 | **.033** |
| VVS lower, NatV1 (reference) | 0.037 | 0.030 | 132.0 | 1.246 | .215 |
| Resting MT | 0.047 | 0.021 | 30.5 | 2.262 | **.031** |
| MADRS | −0.063 | 0.021 | 29.5 | −3.017 | **.005** |
| DVARS | 0.016 | 0.016 | 73.0 | 0.983 | .329 |
| DVS higher, ModV vs. NatV1 | −0.126 | 0.032 | 25520 | −3.972 | **<.001** |
| DVS lower, ModV vs. NatV1 | 0.060 | 0.028 | 25520 | 2.116 | **.034** |
| VVS higher, ModV vs. NatV1 | 0.135 | 0.028 | 25550 | 4.851 | **<.001** |
| VVS lower, ModV vs. NatV1 | −0.224 | 0.033 | 25580 | −6.714 | **<.001** |
| DVS higher, NatV2 vs. NatV1 | −0.047 | 0.032 | 25580 | −1.455 | .146 |
| DVS lower, NatV2 vs. NatV1 | −0.118 | 0.029 | 25510 | −4.031 | **<.001** |

| | | | | | |
|---|---|---|---|---|---|
| VVS higher, NatV2 vs. NatV1 | 0.239 | 0.028 | 25580 | 8.639 | **<.001** |
| VVS lower, NatV2 vs. NatV1 | 0.079 | 0.032 | 25570 | 2.453 | **.014** |

Bold *p* values indicate statistical significance ($p < .05$). df, Satterthwaite degrees of freedom; DVARS, head motion; DVS, dorsal visual stream; MADRS, Montgomery–Åsberg Depression Rating Scale; ModV, Visual Attention Modification run; MT, motor threshold; NatV1, first Natural Face-Viewing run; NatV2, second Natural Face-Viewing run; VVS, ventral visual stream.

***Supplementary Table S4.*** **Pairwise comparisons of dynamic effective connectivity between runs within each visual stream level following iTBS**

| Visual Stream Level | Contrast | Estimate | SE | *z* | *p* (Holm) |
|---|---|---|---|---|---|
| DVS higher | NatV1 vs. ModV | −0.053 | 0.030 | −1.75 | .161 |
| | NatV1 vs. NatV2 | −0.079 | 0.030 | −2.63 | **.025** |
| | ModV vs. NatV2 | −0.026 | 0.030 | −0.87 | .383 |
| DVS lower | NatV1 vs. ModV | −0.002 | 0.028 | −0.08 | .943 |
| | NatV1 vs. NatV2 | 0.027 | 0.029 | 0.93 | .943 |
| | ModV vs. NatV2 | 0.029 | 0.029 | 1.01 | .943 |
| VVS higher | NatV1 vs. ModV | −0.011 | 0.026 | −0.43 | .665 |
| | NatV1 vs. NatV2 | 0.025 | 0.026 | 0.98 | .657 |
| | ModV vs. NatV2 | 0.036 | 0.026 | 1.40 | .485 |
| VVS lower | NatV1 vs. ModV | −0.086 | 0.031 | −2.74 | **.012** |
| | NatV1 vs. NatV2 | −0.110 | 0.031 | −3.49 | **.001** |
| | ModV vs. NatV2 | −0.024 | 0.031 | −0.77 | .439 |

Contrasts represent differences in estimated marginal means on the standardized connectivity scale (first run minus second run). Tests were based on asymptotic ($z$) statistics, and *p* values were Holm-adjusted within each visual stream level across the three run comparisons. Bold *p* values indicate statistical significance ($p < .05$). DVS, dorsal visual stream; ModV, Visual Attention Modification run; NatV1, first Natural Face-Viewing run; NatV2, second Natural Face-Viewing run; VVS, ventral visual stream.

***Supplementary Table S5.* Pairwise comparisons of dynamic effective connectivity between runs within each visual stream level following cTBS**

| Visual Stream Level | Contrast | Estimate | SE | *z* | *p* (Holm) |
|---|---|---|---|---|---|
| DVS higher | NatV1 vs. ModV | 0.126 | 0.032 | 3.97 | **< .001** |
| | NatV1 vs. NatV2 | 0.047 | 0.032 | 1.46 | .146 |
| | ModV vs. NatV2 | −0.079 | 0.032 | −2.52 | **.023** |
| DVS lower | NatV1 vs. ModV | −0.060 | 0.028 | −2.12 | **.034** |
| | NatV1 vs. NatV2 | 0.118 | 0.029 | 4.03 | **< .001** |
| | ModV vs. NatV2 | 0.177 | 0.029 | 6.07 | **< .001** |
| VVS higher | NatV1 vs. ModV | −0.135 | 0.028 | −4.85 | **< .001** |
| | NatV1 vs. NatV2 | −0.239 | 0.028 | −8.64 | **< .001** |
| | ModV vs. NatV2 | −0.105 | 0.027 | −3.90 | **< .001** |
| VVS lower | NatV1 vs. ModV | 0.224 | 0.033 | 6.71 | **< .001** |
| | NatV1 vs. NatV2 | −0.079 | 0.032 | −2.45 | **.014** |
| | ModV vs. NatV2 | −0.303 | 0.034 | −9.01 | **< .001** |

Contrasts represent differences in estimated marginal means on the standardized connectivity scale (first run minus second run). Tests were based on asymptotic ($z$) statistics, and $p$ values were Holm-adjusted within each visual stream level across the three run comparisons. Bold $p$ values indicate statistical significance ($p < .05$). DVS, dorsal visual stream; ModV, Visual Attention Modification run; NatV1, first Natural Face-Viewing run; NatV2, second Natural Face-Viewing run; VVS, ventral visual stream.

### 5. Exploratory Post Hoc Analyses of the Face Inversion Effect

As a post hoc exploratory analysis, we assessed whether stimulation-related changes in the face inversion effect exceeded expected practice effects observed in a separate no-stimulation dataset (see Supplementary Table S6). To minimize potential carryover effects from crossover stimulation exposure, analyses were restricted to participants completing their first stimulation condition only (iTBS-first: $n = 20$; cTBS-first: $n = 19$) and compared against participants completing the task twice without stimulation ($n = 37$). Separate 2 × 2 mixed ANOVAs were conducted for each stimulation condition and duration, with stimulation condition (stimulation vs. no stimulation) as a between-subjects factor and session (pre vs. post) as a within-subjects factor.

***Supplementary Table S6.*** **Sample characteristics of the no-stimulation face inversion effect dataset**

| | **BDD** | **Subclinical BDD** |
|---|---|---|
| **Demographics** | | |
| Total number of participants (female/male) | 25 (17/8) | 12 (9/3) |
| Age, years | 26.16 ± 5.07 | 26.53 ± 5.38 |
| **Symptoms severity** | | |
| DCQ | 15.88 ± 2.42 | 11.58 ± 3.58 |
| BDD-YBOCS | 27.48 ± 4.67 | 17.92 ± 3.29 |
| BABS | 14.28 ± 3.51 | 12.42 ± 4.66 |
| MADRS | 12.32 ± 9.73 | 7.58 ± 6.26 |
| HAM-A | 9.92 ± 8.22 | 5.33 ± 4.92 |
| **Psychiatric comorbidities** | | |
| Major depressive episode | 7 | 0 |
| Persistent depressive disorder | 3 | 0 |
| Obsessive compulsive disorder | 2 | 0 |
| Generalized anxiety disorder | 2 | 0 |
| Social anxiety disorder | 3 | 0 |
| PTSD | 0 | 0 |
| Panic disorder | 1 | 0 |
| No DSM comorbid disorder | 12 | 12 |

Values are presented as mean ± SD; psychiatric comorbidities are presented as counts. BABS, Brown Assessment of Beliefs Scale; BDD, body dysmorphic disorder; BDD-YBOCS, Yale–Brown Obsessive Compulsive Scale modified for BDD; DCQ, Dysmorphic Concern Questionnaire; DSM, Diagnostic and Statistical Manual of Mental Disorders; HAM-A, Hamilton Anxiety Rating Scale; MADRS, Montgomery–Åsberg Depression Rating Scale; PTSD, post-traumatic stress disorder.

*iTBS vs. No-Stimulation:* For short-duration trials, comparison of the iTBS-first subgroup with the no-stimulation group revealed no significant main effect of stimulation condition ($F_{1,55}$ = 0.02, $p$ = .901), no main effect of session ($F_{1,55}$ = 0.74, $p$ = .392), and no stimulation × session interaction ($F_{1,55}$ = 0.21, $p$ = .652). Within-condition contrasts indicated no significant pre- to post-session change in either the no-stimulation group (mean Δ = .028, $p$ = .272) or the iTBS group (mean Δ = −.009, $p$ = .801).

For long-duration trials, comparison of the iTBS-first subgroup with the no-stimulation group similarly revealed no significant main effect of stimulation condition ($F_{1,55} < 0.01$, $p$ = .968), no main effect of session ($F_{1,55} = 0.62$, $p$ = .435), and no stimulation × session interaction ($F_{1,55} = 0.04$, $p$ = .841). Neither the no-stimulation group (mean Δ = .015, $p$ = .624) nor the iTBS group (mean Δ = −.025, $p$ = .542) demonstrated significant pre- to post-session changes.

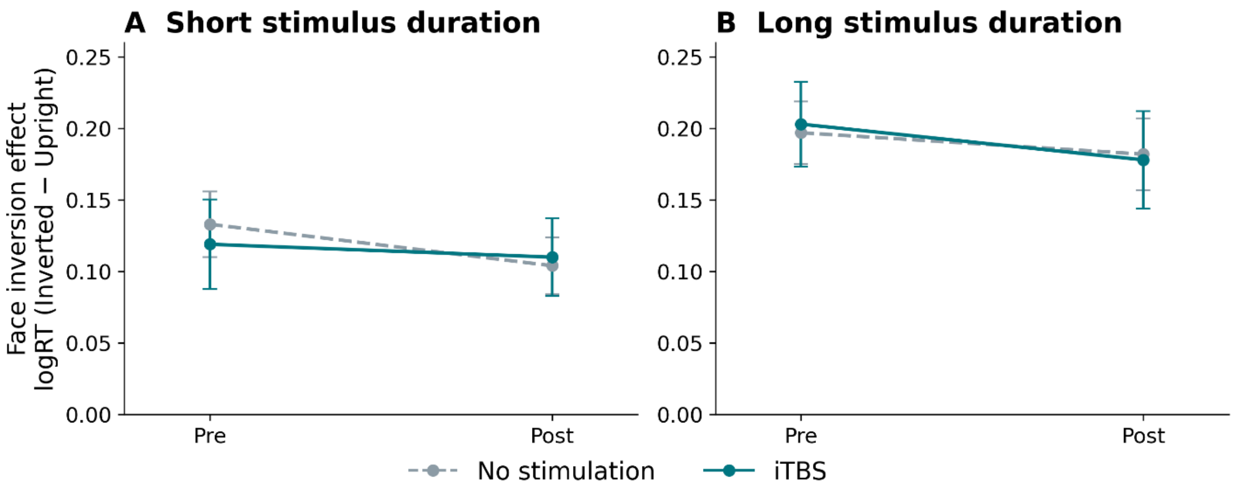


*Supplementary Figure S1*. iTBS versus no-stimulation changes in the face inversion effect. Model-estimated marginal means (logRT, Inverted − Upright) by session; error bars are standard errors. **(A)** short-duration trials; **(B)** long-duration trials. No stimulation $n$ = 37; iTBS-first $n$ = 20.

*cTBS vs. No-Stimulation:* For short-duration trials, comparison of the cTBS-first subgroup with the no-stimulation group also revealed no significant main effect of stimulation condition ($F_{1,54}$ = 0.69, $p$ = .409), no main effect of session ($F_{1,54} = 0.66$, $p$ = .419), and no stimulation × session interaction ($F_{1,54} = 0.15$, $p$ = .704). There were no significant pre- to post-session changes within either the no-stimulation group (mean Δ = −.028, $p$ = .309) or the cTBS group (mean Δ = −.010, $p$ = .792).

For long-duration trials, comparison of the cTBS-first subgroup with the no-stimulation group revealed no significant main effect of stimulation condition ($F_{1,54} = 0.65$, $p$ = .424) and no significant stimulation × session interaction ($F_{1,54} = 1.99$, $p$ = .164). There was a significant main effect of session ($F_{1,54} = 4.10$, $p$ = .048). Within-condition contrasts indicated no significant pre- to post-session change in the no-stimulation group (mean Δ = −.015, $p$ = .600), whereas the cTBS group demonstrated a significant reduction in the face inversion effect from pre- to post-session (mean Δ = .084, $p$ = .039). However, because the stimulation × session interaction was not significant, this effect should be interpreted cautiously.

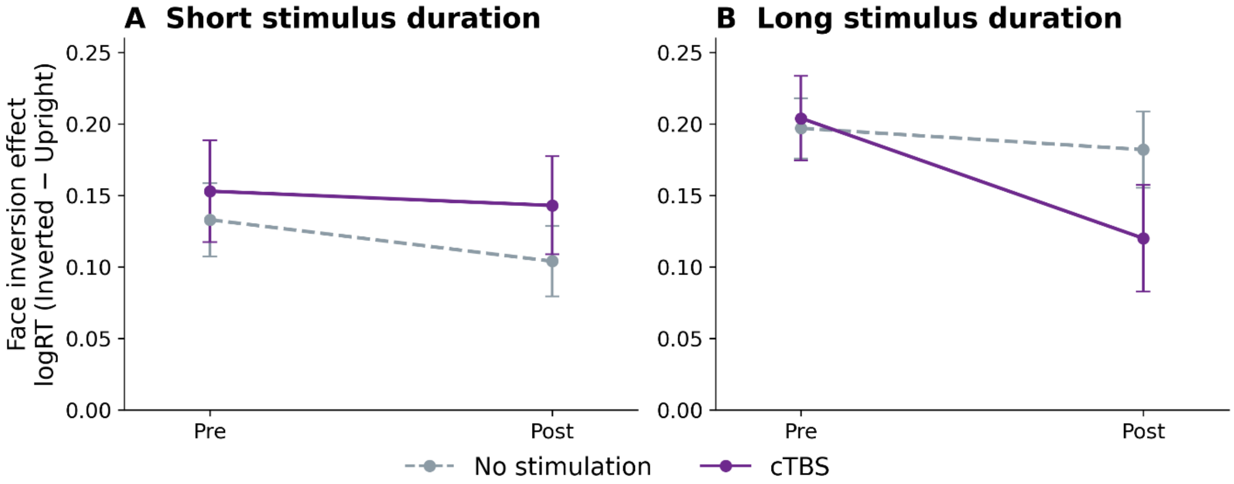


*Supplementary Figure S2.* cTBS versus no-stimulation changes in the face inversion effect. Model-estimated marginal means (logRT, Inverted − Upright) by session; error bars are standard errors. **(A)** short-duration trials; **(B)** long-duration trials. No stimulation *n* = 37; cTBS-first *n* = 19.

### 6. Self-Reported Mood, Anxiety, Distress, and Appearance Concern

The Body Image States Scale (BISS) (20) was collected to assess momentary appearance satisfaction prior to and following administration of both TBS and ModV, while the State-Trait Anxiety Inventory (STAI-S) (21) was collected once prior to administration of TBS and once following ModV to assess momentary anxiety states. The Face Concern Visual Analog Scale (VAS) and Brief Subjective Distress Ratings were also completed following ModV to assess participants perceived level of distress after viewing photographs of their own face.

*Body Image State and Anxiety*. Scores on the BISS decreased modestly from pre- to post-stimulation across both TBS conditions (Supplementary Table S7). A significant main effect of session was observed for BISS scores ($F_{1,36}$ = 5.37, $p$ = .026), indicating lower post-stimulation scores regardless of stimulation type. However, there was no effect of TBS condition on BISS change scores ($F_{1,36}$ = 0.85, $p$ = .363).

State anxiety, measured using the State-Trait Anxiety Inventory (STAI-State), did not significantly change across sessions ($F_{1,36}$ = 0.95, $p$ = .337) and there was no effect of TBS condition on anxiety change scores ($F_{1,36}$ = 0.60, $p$ = .444). A trend-level effect of participant type was observed ($F_{1,36}$ = 4.01, $p$ = .053).

***Supplementary Table S7*. Body image state and anxiety scores before and after stimulation**

| **Measure** | **Effect** | ***F*** | ***p*** |
|---|---|---|---|
| BISS Total Score | TBS Condition | 0.85 | .363 |
| | Session (Pre vs. Post) | 5.37 | **.026** |

| | | | |
|---|---|---|---|
| | Participant Type | 1.22 | .277 |
| STAI-State Score | TBS Condition | 0.60 | .444 |
| | Session (Pre vs. Post) | 0.95 | .337 |
| | Participant Type | 4.01 | .053 |

Bold *p* values indicate statistical significance ($p < .05$). BISS, Body Image States Scale; STAI-State, State-Trait Anxiety Inventory—State; TBS, theta burst stimulation.

*Distress and Face Concern Ratings*. Participants provided ratings of subjective distress and face concern during the experimental sessions (Supplementary Table S8). Distress ratings did not differ between TBS conditions ($F_{1,37} = 0.18$, $p = .677$) and showed only a trend-level effect of session ($F_{1,37} = 3.81$, $p = .059$). Similarly, face concern ratings did not differ between stimulation conditions, ($F_{1,37} = 0.04$, $p = .849$), although a significant effect of session was observed ($F_{1,37} = 7.17$, $p = .011$). Significant effects of participant type were observed for both distress ratings ($F_{1,37} = 11.51$, $p = .002$), and face concern ratings ($F_{1,37} = 10.25$, $p = .003$) (see Supplementary Table S9).

***Supplementary Table S8*. Distress and face concern ratings during experimental sessions**

| **Measure** | **TBS Condition** | **N** | **Mean (SD)** |
|---|---|---|---|
| Distress Rating | iTBS | 39 | 5.21 (2.61) |
| Distress Rating | cTBS | 38 | 4.97 (2.75) |
| Face Concern Rating | iTBS | 39 | 51.31 (22.18) |
| Face Concern Rating | cTBS | 38 | 51.34 (26.73) |

cTBS, continuous theta burst stimulation; iTBS, intermittent theta burst stimulation.

***Supplementary Table S9*. Mixed-effects model results for distress and face concern ratings**

| **Measure** | **Effect** | ***F*** | ***p*** |
|---|---|---|---|
| Distress Rating | TBS Condition | 0.18 | .677 |
| | Session | 3.81 | .059 |
| | Participant Type | 11.51 | **.002** |
| Face Concern Rating | TBS Condition | 0.04 | .849 |
| | Session | 7.17 | **.011** |
| | Participant Type | 10.25 | **.003** |

Bold $p$ values indicate statistical significance ($p$ < .05). TBS, theta burst stimulation.
